# Combining fine-scale social contact data with epidemic modelling reveals interactions between contact tracing, quarantine, testing and physical distancing for controlling COVID-19

**DOI:** 10.1101/2020.05.26.20113720

**Authors:** Josh A. Firth, Joel Hellewell, Petra Klepac, Stephen Kissler, CMMID COVID-19 working group, Adam J. Kucharski, Lewis G. Spurgin

## Abstract

Case isolation and contact tracing can contribute to the control of COVID-19 outbreaks^1,2^. However, it remains unclear how real-world networks could influence the effectiveness and efficiency of such approaches. To address this issue, we simulated control strategies for SARS-CoV-2 in a real-world social network generated from high resolution GPS data^3,4^. We found that tracing contacts-of-contacts reduced the size of simulated outbreaks more than tracing of only contacts, but resulted in almost half of the local population being quarantined at a single point in time. Testing and releasing non-infectious individuals led to increases in outbreak size, suggesting that contact tracing and quarantine may be most effective when it acts as a ‘local lockdown’ when contact rates are high. Finally, we estimated that combining physical distancing with contact tracing could enable epidemic control while reducing the number of quarantined individuals. Our approach highlights the importance of network structure and social dynamics in evaluating the potential impact of SARS-CoV-2 control.

## Main

Non-pharmaceutical interventions (NPIs) are central to reducing SARS-CoV-2 transmission ^5–8^. Such responses generally include: case isolation, tracing and quarantining of contacts, use of PPE and hygiene measures, and policies designed to encourage physical distancing (including closures of schools and workplaces, banning of large public events and restrictions on travel). Due to the varying economic and social costs of these interventions, there is a clear need for sustainable strategies that limit SARS-CoV-2 transmission while reducing disruption as far as possible.

Isolation of symptomatic cases, and quarantine of their contacts (e.g. household members), is a common public health strategy for reducing disease spread^1,2,8^ This approach has been used as part of SARS-CoV-2 control strategies^9^. However, the relatively high reproduction number of the SARS-CoV2 virus in early outbreak stages^10,11,^ alongside likely high contribution to transmission from presymptomatic and asymptomatic individuals^12^, means that manual tracing of contacts alone may not be a sufficient containment strategy under a range of outbreak scenarios^13^. As countries relax lockdowns and other more stringent physical distancing measures, combining the isolation of symptomatic individuals and quarantine of contacts identified through fine-scale tracing is likely to play a major role in many national strategies for targeted SARS-CoV-2 control^14^

It is possible to assess the potential effectiveness of contact tracing by simultaneously modelling disease spread and contact tracing strategies through social systems of individuals^15^ These systems are usually simulated through parameterisation with simple social behaviours (e.g. the distribution of the number of physical contacts per individual). Further still, social systems may be simulated as networks that can be parameterised according to assumptions regarding different contexts (for example, with different simulated networks for households, schools and workplaces), or using estimated contact rates of different age groups^16^ However, much less is known about how different types of real-world social behaviour and the hidden structure found in real-life networks may affect both patterns of disease transmission and efficacy of contact tracing under different scenarios^17,18^. Examining contagion dynamics and control strategies using a ‘real-world’ network allows for a more realistic simulation of SARS-CoV-2 outbreak and contact tracing dynamics.

Here we develop an epidemic model which simulates COVID-19 outbreaks across a real-world network, and we assess the impact of a range of testing and contact tracing strategies for controlling these outbreaks (Table 1). We then simulate physical distancing strategies and quantify how the interaction between physical distancing, contact tracing and testing affects outbreak dynamics.

**Table 1.** Policy summary

|  |  |
| --- | --- |
| Background | Understanding how isolation, contact tracing and other non-pharmaceutical interventions can combine effectively and efficiently is crucial to COVID-19 control. Such interventions are likely to depend on contact patterns within a population. We developed an epidemic model that simulates COVID-19 outbreaks in a real-world network, and assess the impact of a range of testing, isolation, quarantine and contact tracing strategies for controlling these outbreaks. |
| Main findings and limitations | We found that isolation, contact tracing and quarantine reduced simulated outbreak size in our local-scale network. Tracing and quarantining contacts of contacts was more effective, but required large numbers of individuals are required to be quarantined. This strategy is therefore often similar to introducing a 'local lockdown'. Testing and releasing quarantined individuals reduced the numbers quarantined, but also the effectiveness of control measures. Combining physical distancing with contact tracing resulted in reduced outbreak size, with fewer individuals required to quarantine. A major limitation of this study is that it is based on pre-COVID-19 data from a sample of individuals from a single town; more data are therefore needed to fully understand potential outbreak dynamics in other settings. |
| Policy implications | Our findings suggest that effective contact tracing measures may require large numbers of people in a community to be quarantined, with individual-level tracing resulting in scenarios equivalent to broader localised lockdowns. Targeted tracing and quarantine strategies may therefore be more efficient when combined with other control measures such as physical distancing. |

We used a publicly available dataset on human social interactions collected specifically for modelling infectious disease dynamics as part of the British Broadcasting Corporation (BBC) documentary “Contagion! The BBC Four Pandemic”. The high-resolution data collection focused on residents of the town of Haslemere, where the first evidence of UK-acquired infection with SARS-CoV-2 would later be reported in late February 2020^19^. Previous analyses of this dataset have shown that it is structurally relevant to modelling disease spread, and hence holds substantial potential for understanding and controlling real-world diseases^4^. Here, we defined dyadic contacts on a day-by-day basis as at least one daily 5 min period with a distance of 4 m (see Methods), which gave 1616 daily contact events and 1257 unique social links between 468 individuals. The social network defined in this way was strongly correlated (*r* >0.85 in all cases) with social networks based on other contact distances (from 1-7 m contact ranges; Extended Data Fig. 1). Similarly, social networks created using different time-periods for weighting the dyadic contacts (Extended Data Fig. 2) were also strongly related to the weighting used here (i.e. number of days seen together). As such, this social network quantification gives a representative indication of daily contact propensities within the relevant transmission range between individuals (see Methods) and also captures various aspects of the patterns and structure presented by different quantifications of this social system.

**Figure 1.**
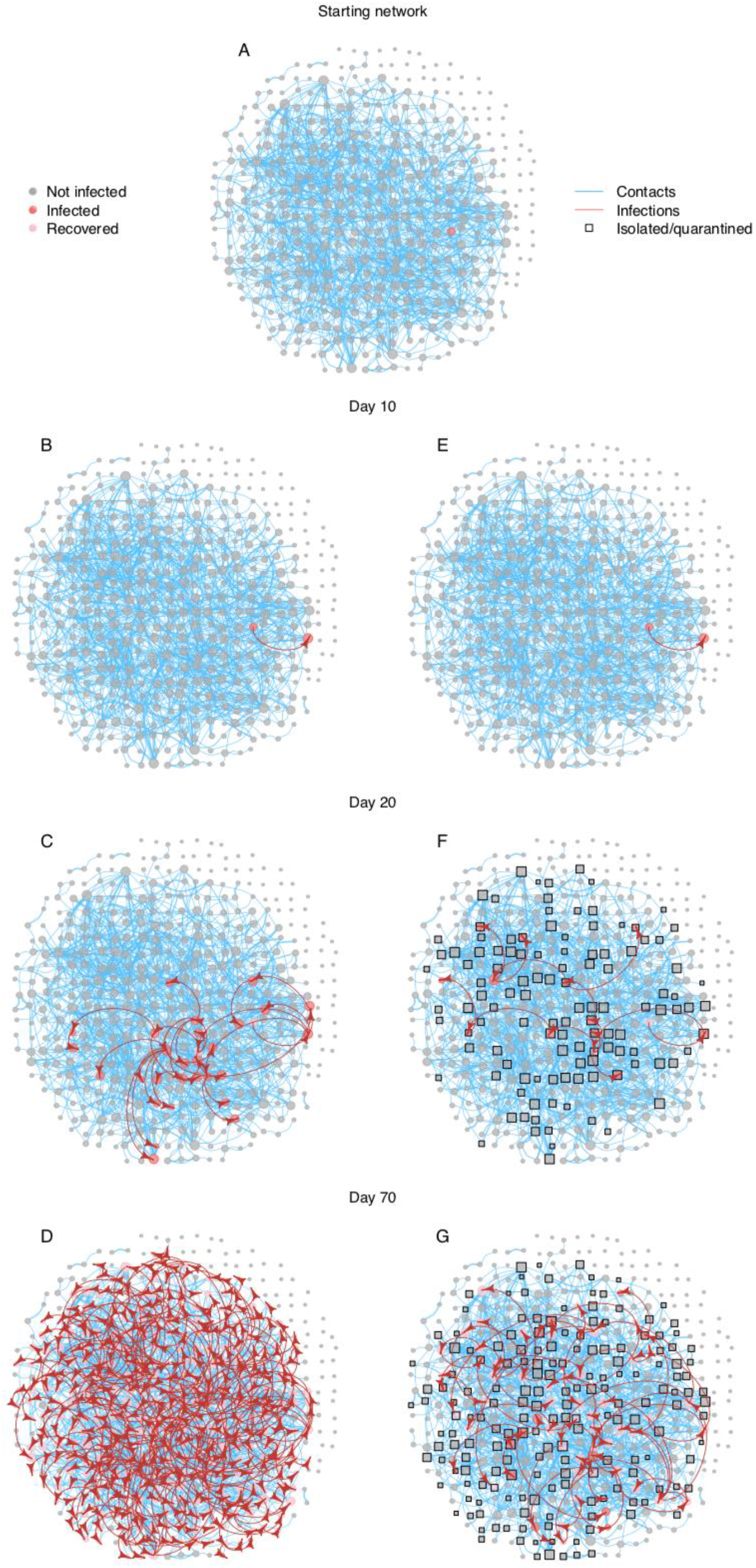
Illustration of the Haslemere network with epidemic simulation predictions. **A** The social network of 468 individuals (grey nodes) with 1257 social links (blue edges) weighted by 1616 daily contacts (edge thickness) and a single starting infector (red). Subsequent panels show progression of the COVID-19 epidemic under the no intervention (**B**,**C**,**D**) and the secondary contact tracing (**E**,**F**,**G**) scenarios. Red arrows show an infection route, and squares show isolated/quarantined individuals.

**Figure 2.**
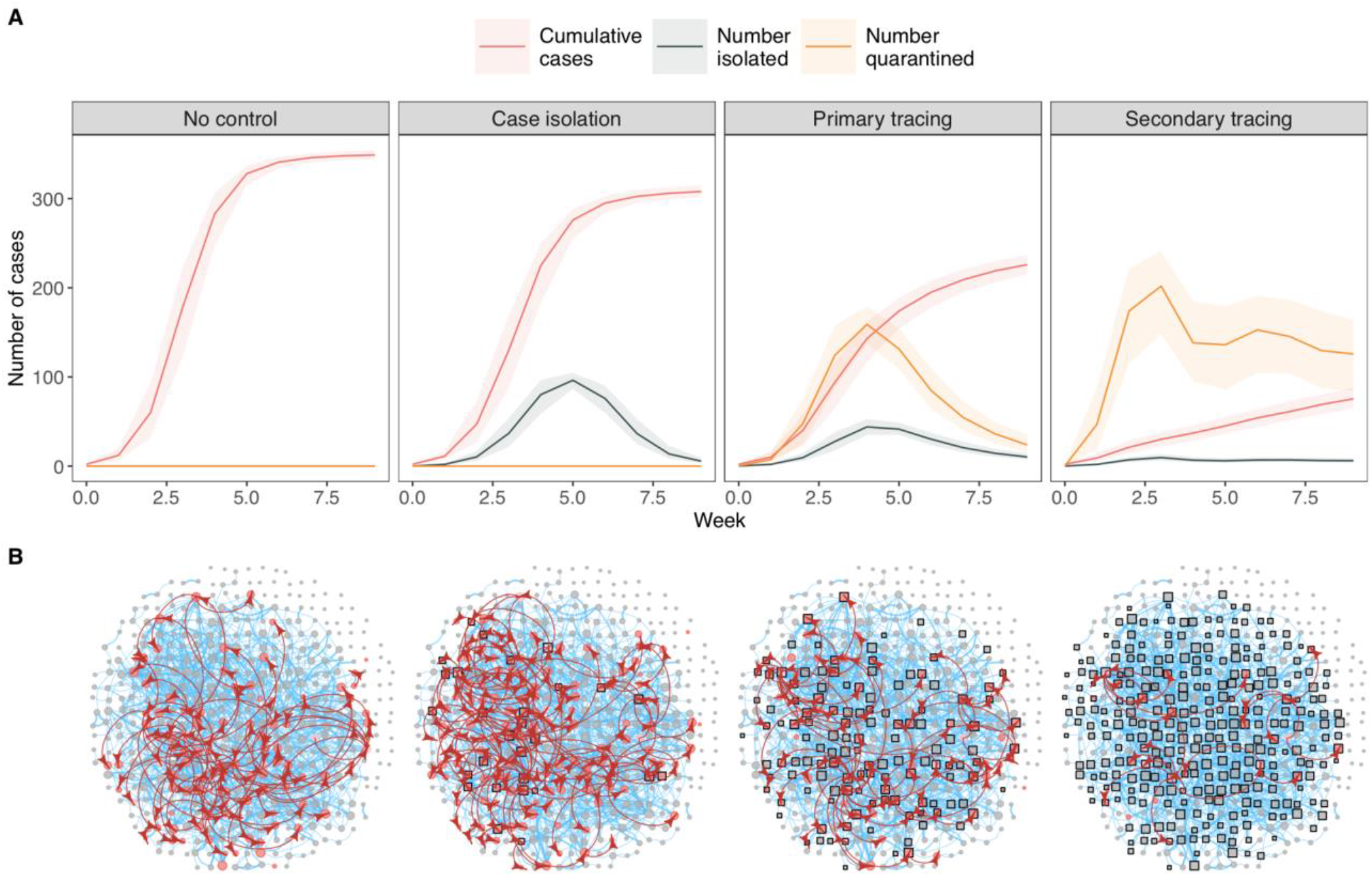
Epidemic model predictions of outbreak size and number of people isolated/quarantined under different non-pharmaceutical intervention scenarios in the Haslemere network. **A** cumulative number of cases, number of people isolated, and number of people quarantined at a given point in time under each scenario. Lines and shaded areas represent median and interquartile range from 1000 simulations. **B** Example networks from a single simulation of each scenario at day 20 of the outbreak. See figure 1 for network details.

Example outbreaks across the Haslemere social network under different control scenarios are displayed in Fig. 1, with a full animated visualisation in Supplementary Video 1, and a Shiny app is available to run individual outbreak simulations (see data sharing). Across all simulations, our epidemic model showed that uncontrolled outbreaks in the Haslemere network stemming from a single infected individual resulted in a median of 75% (IQR = 74%-76%) of the population infected after 70 days (Fig. 2). Isolation when symptomatic resulted in 66% (65%-67%) of the population infected, while primary contact tracing resulted in 48% (46%-50%) infected. Secondary contact tracing resulted in the smallest percentage (16%, 14%-19%) of the population infected after 70 days. The number of quarantined individuals was very high under both primary and secondary contact tracing, with a median of 43% (IQR = 32%-52%) of the population quarantined during the outbreak peak with the latter (Fig. 2). Examining temporal dynamics showed that outbreak peaks typically occurred within the first 1-3 weeks, and that interventions reduced the overall size of the outbreaks as well as their growth rate (Fig. 2). The number of people required to isolate or quarantine followed a similar trajectory to the number of cases, although under secondary contact tracing, substantial proportions of the population (27%, 18%-35%) were quarantined even at the end of the simulations (Fig. 2). This is in line with a large-scale recent simulation model of app-based contact tracing in the UK^20^, which suggested that contact tracing could be highly effective, but also that it required large numbers of people to be quarantined. Further, in our (optimistic) default parameter settings we assumed that 10% of contact tracing attempts were missed. This, combined with the very large number of quarantined cases under secondary contact tracing (Fig. 2), suggests that a majority of the population could receive a notification that they should quarantine within the first 2-3 weeks of an outbreak.

Sensitivity analysis of the efficacy of contact tracing under the epidemic model is presented in Extended Data Figs 3-6. As expected, outbreak size decreased with the percentage of contacts traced in all scenarios, and increased with the reproduction number, the proportion of asymptomatic cases, the proportion of pre-onset transmission, the delay between onset/tracing and isolation/quarantine, and the number of initial cases (Extended Data Figs 3-6). Outbreak dynamics were strongly affected by outside infection rate across all intervention scenarios, as were the number of isolated and quarantined cases (Extended Data Fig. 6). Our model therefore corroborates with models using simulated social systems and showing that, for a disease such as COVID-19 with high levels of transmission from asymptomatic and presymptomatic individuals, contact tracing is likely to be most effective when the proportion of traced contacts is high, when the delay from notification to quarantine is short^13^, and, most importantly, when the number of starting cases and rate of movement into the network are low. Importantly, however, the tradeoff between the number of cases and the number of quarantined cases was found across the entirety of the parameter space (Extended Data Figs 3-6). Further, increasing the network density through increasing the distance threshold for defining contacts led to broadly similar results across intervention scenarios, albeit with larger numbers of quarantined cases required for outbreak control via contact tracing (Extended Data Fig. 7). Therefore, while more real-world networks are needed to demonstrate how well these results apply to other locations and settings, our results are robust to a range of epidemiological and network parameters.

**Figure 3.**
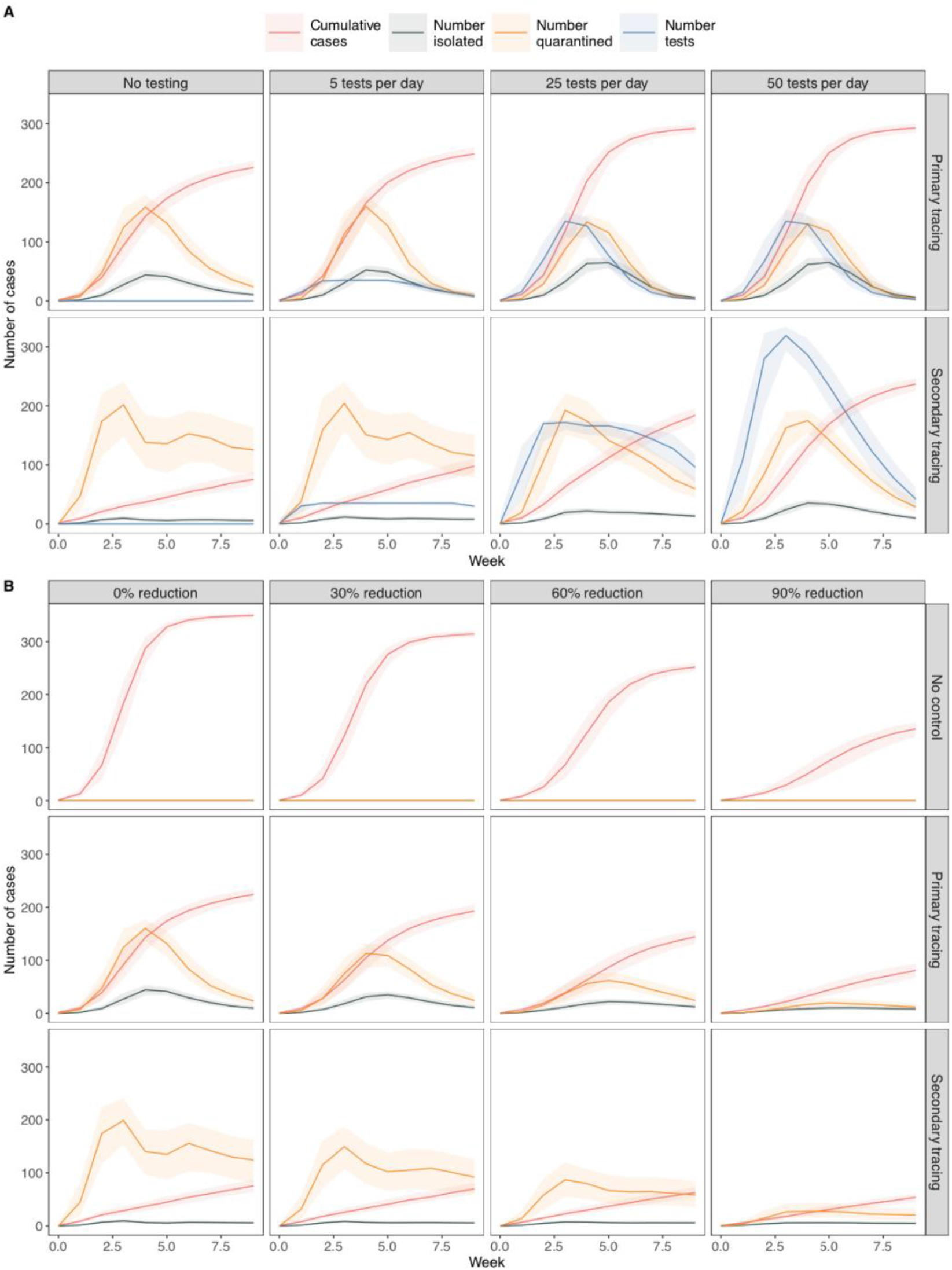
**A** Epidemic model simulations of outbreak size and number of people isolated and quarantined under **A** different levels of testing and **B** physical distancing in the Haslemere network. In **A**, Tests are plotted per week rather than per day for visualisation purposes. In **B** The percentage reduction refers to the number of ‘weak links’ removed from the networks (see methods). Lines and shaded areas represent median and interquartile range from 1000 simulations.

The number of quarantined cases can be reduced through mass testing and release of individuals who return a negative result. Conversely, if contact rates in the population are high, large-scale test and release strategies could provide greater opportunity for transmission and decrease the effectiveness of contact tracing. We therefore assessed how the testing and releasing of isolated and quarantined subjects might affect the numbers of cases and time spent in isolation and quarantine, using false positive and false negative rates estimated from empirical data^21,22^ (Supplementary Table 1). We estimated that increasing the testing capacity (and therefore testing and releasing more quarantined cases) led to substantial increases in outbreak size, especially under secondary contact tracing (median = 50%, IQR = 48%-52%; Fig. 3A). This result occurred despite an optimistic false negative rate of 10%, suggesting that the increase in outbreak size with high testing rates is a result of increased transmission within the network, rather than through releasing infected cases *per se*. Therefore, secondary tracing may effectively act as a ‘local lockdown’ rather than a targeted intervention strategy. High levels of testing did not lead to large reductions in the number of quarantined cases under secondary contact tracing scenarios, and the number of tests required to reduce the numbers of quarantined cases were large, with 68% (63%-71%) of the population requiring tests in a single week during outbreak peaks (Fig. 3A). We cannot be sure to what extent our results will represent larger populations, but the tripartite relationship between the number of cases, the number of quarantined contacts and the number of tests required will apply in the majority of scenarios in which rates of social interaction are high.

A very high notification and quarantine rate for any contact tracing system may have consequences for adherence. Our model is optimistic in its assumption that individuals isolate independently of previous notifications or isolations, and highly optimistic in its assumption of 100% adherence to quarantine among traced contacts. In reality a high notification and quarantine rate may result in individuals being less likely to undertake quarantine in the future, which in turn will impact outbreak dynamics. There is a need for more evidence and models to better understand these behavioural dynamics, in order to develop sustainable intervention strategies^23^ One suggested solution to reduced adherence to quarantine is through (digital) targeted quarantine requests to the individuals at highest risk of infection, or to those most likely to spread to others^24^ To what extent these interventions will be needed, and how well they will work, is not yet clear; however, our study provides a methodological template for network-based research into SARS-CoV2 and its potential control strategies.

Combining contact tracing with other physical distancing measures may allow for outbreak control while reducing the number of people in quarantine, and the number of tests required. We simulated physical distancing by reducing the number of weak links in the Haslemere network. We aimed to consider low to moderate levels of physical distancing, so used a model whereby the only interactions with ‘rare’ contacts are removed. We found that, across all scenarios, physical distancing led to reductions in the number of overall cases (Fig. 3B). Importantly, increasing physical distancing was associated with lower numbers of quarantined cases, which was reduced to as little as 6% (3%-9%) during outbreak peaks under secondary contact tracing (Fig. 3B). Simulating physical distancing using an alternative approach whereby removed ‘rare’ contacts were reassigned to existing contacts (see methods) yielded similar results to our simpler model, although using this approach, physical distancing led to smaller decreases in outbreak size (Extended Data Fig. 8). We do not have information on household structure within the Haslemere dataset, but our physical distancing scenario is analogous to decreasing the level of non-household contacts. Therefore it may be that combining measures that reduce non-household contact rates with highly effective contact tracing may be a useful tool for control of SARS-CoV-2 spread. However, further work is required to determine exactly what kinds of physical distancing measures would enable effective outbreak control alongside contact tracing. Furthermore, future investigations could also examine how the spread of the disease itself may shape behavioural change interventions (e.g. where large outbreaks spark more severe physical distancing measures), and how this feedback may shape the contagion dynamics and predicted effectiveness of interventions.

Network structure can have substantial effects on epidemic model predictions^25,26^ To investigate this, we used null network models based on the Haslemere data, which maintained the same number of individuals, connections and weights of connections, but shuffled network architecture in different ways (see Methods). The number of cases estimated using the null networks was broadly similar to the real-world network, although this was substantially underestimated in a ‘lattice’ like network (Fig. 4). Importantly, the rate of quarantine varied substantially among the null networks, especially under secondary contact tracing (Fig. 4). These results demonstrate that the use of network-based simulations of SARS-CoV-2 dynamics requires caution, as even if such models had precise information on the number of individuals and amount of social interactions occurring within a system, the assumed architecture of the social network structure alone can shape predictions for both the extent of spread and the usefulness of control strategies. Furthermore, through providing insight into how changes to network structure influences contagion dynamics, the null network simulation approach gives some indication of how this contagion and associated control strategies may operate in different social environments. For instance, different social structures may arise when considering particular social settings (e.g. workplaces, commuting), some of which may be closer to the null networks generated here. Considering this structure will lead to improved predictions of outbreak dynamics.

**Figure 4.**
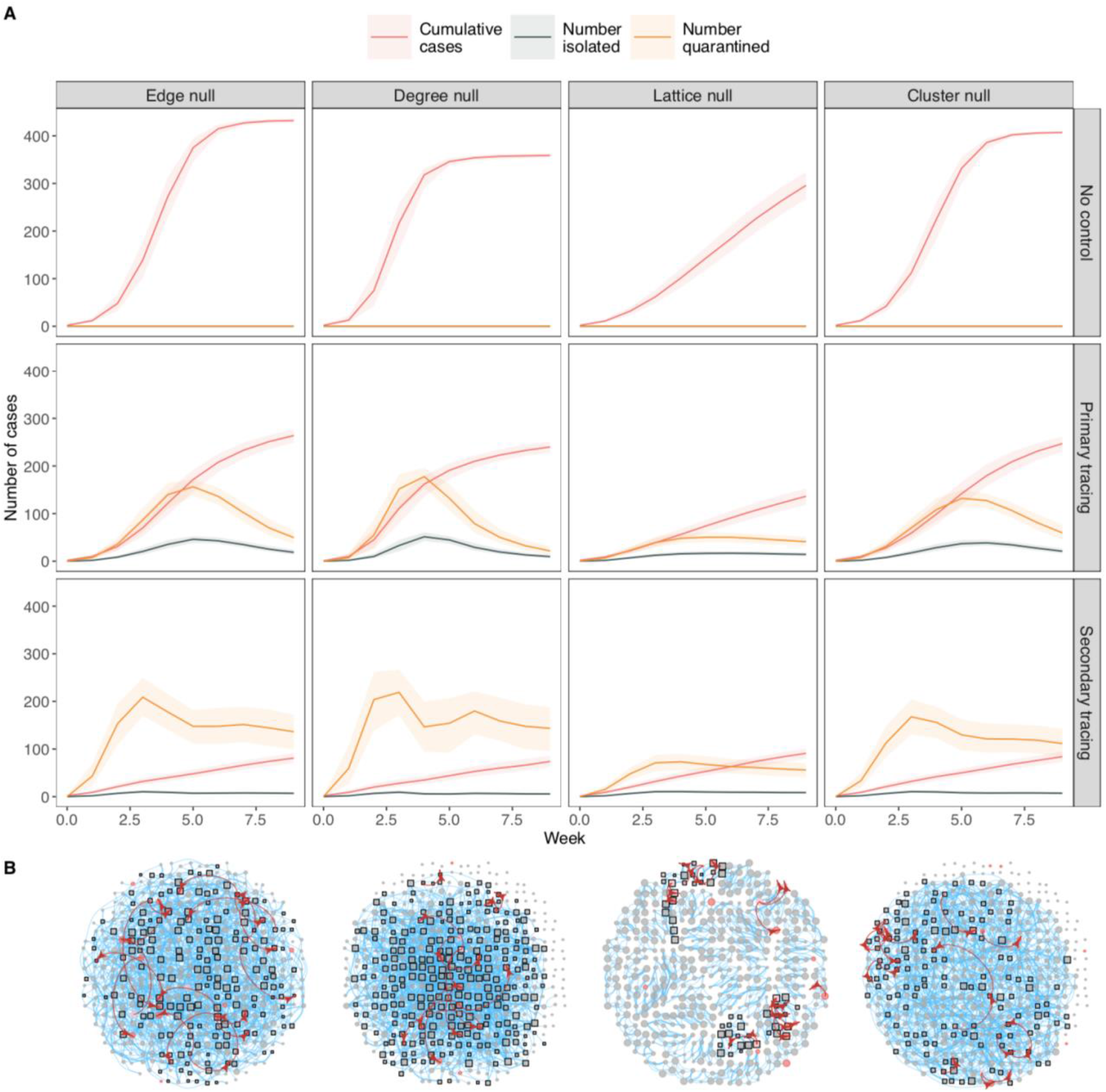
**A** Epidemic model simulations of outbreak size and number of people isolated and quarantined under different null-network permutations based on the Haslemere network (see methods for details). Lines and shaded areas represent median and interquartile range from 1000 simulations. **B** Example networks showing an infection simulation (with secondary contact tracing, after 20 days) on each null network. See Figure 1 for network details.

There are a number of important limitations to our study and the current availability of empirical data in general. Most importantly, this social network is taken from a single, small town and over a short period of time, and we do not know to what extent the social dynamics will be applicable to larger cities and other contexts and over long periods. Therefore, future large-scale efforts in gathering data on dynamic fine-scale social behaviour over long periods of time (ideally over the entire contagion period) in major cities would be of great benefit for assessing the relative uses of SARS-CoV-2 control strategies, and for understanding how and why interventions implemented in some cities have been relatively more successful than others^27^ Indeed, the epidemic network-based model provided here can be applied generally to more extensive social networks if such data becomes available in the future. Further, the Haslemere data, while rich, does not sample the entire population of Haslemere, and children under the age of 13 were not included in the experiment, which could potentially have an impact on outbreak and social tracking dynamics. Again, such issues are also likely to be prevalent across real-world contact-tracing attempts, as the ability to track children will be limited, particularly with app-based approaches that require a smartphone. It is encouraging that our results broadly align with other, larger-scale simulations of contact tracing which explicitly model these limitations, but lack the fine-scale social tracking data^20^ Therefore, by supplying a general framework for simulating the spread of COVID-19 on real-world networks, we hope to promote integration of multiple real-world social tracking datasets with epidemic modelling, which may provide a promising way forward for optimising contact tracing strategies and other non-pharmaceutical interventions.

## Data Availability

This study used the raw data previously published in Kissler et al. 2018 (made available with full description here: https://www.biorxiv.org/content/10.1101/479154v1)
The code and data used to produce the simulations is available as an R package at: https://github.com/biouea/covidhm
A shiny app which runs individual outbreak simulations is available at:
https://biouea.shinyapps.io/covidhm_shiny/

https://github.com/biouea/covidhm

## Methods

### Social tracking data

The Haslemere dataset was generated and described as part of previous work, which gives detailed description of the characteristics of this dataset and town^3,4^ Briefly, the data were collected during the 2017/18 *BBC Pandemic* project conducted in Haslemere, Surrey, UK. The project involved a massive citizen-science experiment to collect social contact and movement data using a custom-made phone app, and was designed to generate data relevant to understanding directly transmitted infectious disease^3,4^ Of the 1272 individuals within Haslemere that downloaded the app, 468 individuals had sufficient data points at a resolution of 1m over three full days within the focal area for further analysis^3^ All 468 focal individuals were known to have spent >6hrs within 51.0132;-0.7731SW : 51.1195,-0.6432NE (within Postcode GU27), but the dataset used here comprises of de-identified proximity data made available as pairwise distances (∼1 m resolution) at 5 min intervals (excluding 11pm-7am)^3^

### Social network construction

In our primary analysis, we defined social contacts as events when the average pairwise distances between individuals within a 5 min time interval (calculated using the Haversine formula for great-circle geographic distance^3^) are 4 m or less. By doing so, we aimed to capture the majority of relevant face-to-face contacts (i.e. those that may result in transmission) over 5 min periods, particularly given the 1 m potential error^3^ on the tracking measurement during these short time intervals. Furthermore, this 4 m threshold is within typical mobile phone Bluetooth ranges for relatively accurate and reliable detections. Therefore, this contact dataset will also be comparable to proximity-based contacts identified through Bluetooth contact tracing apps, which may be preferred to real-location tracking for privacy reasons. We considered the sensitivity of the network to the contact definition by testing six further social networks from contacts defined using different threshold distances spanning the conceivable potential transmission range within the 5 min intervals (1 m to 7 m thresholds). We first measured the correlation of the network structure (i.e. pairwise contacts) across the seven networks using Mantel tests. We also measured the correlation of each individual’s degree (number of contacts), clustering coefficient (number of contacts also connected to one another), betweenness (number of shortest paths between nodes that pass through an individual), and eigenvector centrality (a measure that accounts both for a node’s centrality and that of its neighbours) across the seven networks.

The Haslemere data is a temporal dataset spanning three full days. While the epidemic model we use is dynamic (see below Methods), the contagion process of COVID-19 operates over a longer time period than three days. To be able to meaningfully simulate longer-term outbreak dynamics, we quantified the data as a static social network in which edges indicate the propensities for social contact between nodes. Temporal information is incorporated by weighting the edges using the temporal contact information, instead of using a dynamic network which would require contact data over a much longer period. In the primary analysis, we weighted the edges as the number of unique days a dyad was observed together (but see Supplementary Information for other temporal definitions). Therefore, the weight score indicates the propensity for each dyad to engage in a social contact event on any given day, whereby 0 = no contact, 1 = ‘weak links’ observed on the minority of days (one third), 2 = ‘moderate links’ observed on the majority of days (two thirds), and 3 = ‘strong links’ observed on all days. In this way, the weights of this social network could be included directly, and intuitively, into the dynamic epidemic model (see below). For sensitivity analysis, we also created other weighings for this network, and examined the correlation in dyadic social associations scores (using Mantel tests) with our primary weighting method (described above). Specifically, for the sensitivity analysis, we used edges specified as i) a binary (i.e. unweighted) network across all days, ii) a raw (and ranked) count of 5 min intervals in contact, iii) a transformed weighted count (edge weight transformed as 1 − *e*^*interval count*^, which approximates a scenario where infection risk increases with contact time, but begins to level off after ∼30mins of contact between dyads) and iv) a ‘simple ratio index’ (SRI) weighting that corrects for observation number as SRI score^28^ The SRI score for any two individuals (i.e. A and B) is calculated as:

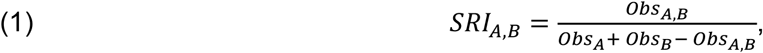

where *Obs* is the number of 5 min observation periods (the intervals since the start of the day) within which an individual is recorded within 4 m of another individual.

### Null network simulation approach

We used null networks^29^ to understand the network properties that shape predictions of COVID-19 spread under different control scenarios. Null networks can also show how contagion may depend on the arrangement of social ties, how it may operate in different social environments, and which simulation approaches may be the most similar to real-world infection dynamics. We created four null network scenarios (Extended Data Fig. 9) with 1000 networks generated under each of these. All of the null network scenarios kept the same number of nodes, edges, and weights of these edges, as the Haslemere network, but were generated under the following nulls: (1) ‘edge null’ (Extended Data Fig. 9A) considered random social associates, allowing the edges of the network to be randomly allocated between all nodes; (2) ‘degree null’ (Extended Data Fig. 9B) considered individual differences in sociality but random social links between dyads, so randomly swapped the edges between nodes but maintained the degree distribution of the real network (and was, therefore, even more conservative than a power-law network simulation aiming to match real differences in sociality); (3) ‘lattice null’ (Extended Data Fig. 9C) considered triadic and tight clique associations, so created a ring-like lattice structure through assigning all edges into a ring-lattice where individuals are connected to their direct neighbours, and their neighbours of the second and third order (i.e. six links per individual), and then we randomly removed excess links (until the observed number of edges was reached); (4) ‘cluster null’ (Extended Data Fig. 9D) considered the observed level of clustering, so created a ring-lattice structure as described above but only between individuals observed as connected (at least 1 social link) in the real network, added remaining links (sampled from 4th order neighbours), and then rewired the edges until the real-world global clustering was observed (∼20% rewiring; Extended Data Fig. 9D). These conservative (and informed) null models allowed connections to be arranged differently within the network but maintained the exact same number of individuals, social connections and weights of these social connections at each simulation.

### Epidemic model

Building on the epidemiological structure of a previous branching-process model^13,^ we developed a full epidemic model to simulate COVID-19 dynamics across the Haslemere network. Full model parameters are given in Supplementary Table 1. For a given network of individuals, an outbreak is seeded by randomly infecting a given number of individuals (default = 1). The model then moves through daily time steps, with opportunities for infection on each day. All newly infected individuals are assigned an ‘onset time’ drawn from a Weibull distribution (mean = 5.8 days) that determines the point of symptom onset (for symptomatic individuals), and the point at which infectiousness is highest (for all individuals)^12^ Each individual is then simultaneously assigned asymptomatic status (whether they will develop symptoms at their onset time), as well as presymptomatic status (whether or not they will infect before their assigned onset time), drawn from Bernoulli distributions with defined probabilities (defaults = 0.4 and 0.2 respectively, see Supplementary Table 1). At the start of each day, individuals are assigned a status of susceptible, infectious or recovered (which would include deaths) based on their exposure time, onset time and recovery time (calculated as onset time plus seven days), and are isolated or quarantined based on their isolation/quarantine time (described below). The model then simulates infection dynamics over 70 days.

Possible infectors are all non-isolated and non-quarantined infectious individuals. Each day, all susceptible, non-isolated, non-quarantined contacts of all infectors within the network are at risk of being infected. The transmission rate for a given pair of contacts is modeled as:

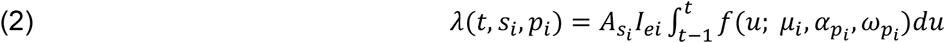

where t is the number of days since the infector *i* was exposed, *s*_*i*_and *p*_*i*_ are the infector’s symptom status (asymptomatic yes/no, and presymptomatic yes/no, respectively). 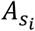 is the scaling factor for the infector’s symptomatic status (Supplementary Table 1) and *I*_*ei*_ is the weighting of the edge in the network (i.e. number of days observed together) between the infector and the susceptible individual. The probability density function 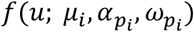 corresponds to the generation time, which is drawn from a skewed normal distribution (see ^13^ for details). Briefly, this uses the infector’s onset time as the location parameter *μ*^*i*^, while the slant parameter 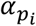 and the scale parameter 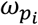 both vary according to the infector’s presymptomatic transmission status (Supplementary Table 1). This enabled us to simulate a predefined rate of presymptomatic transmission, while retaining a correlation structure between onset time and infectiousness, and avoiding a scenario whereby a large number of individuals were highly infectious on the first day of exposure (see Supplementary Table 1 and data sharing for more details).

Using this transmission rate, the probability of infection between a susceptible-infected pair of individuals *t* days after the infector’s exposure time is then modeled as:

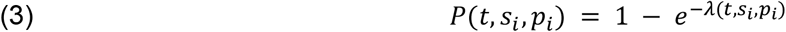

Note that the change in status from “infectious” to “recovered” at seven days after symptom onset does not affect infection dynamics (as transmission rate ≈ 0 seven days after onset time in our model), but is instead used for contact tracing purposes (see below). To test how the above rate of infection related to the reproduction number *R*_0_ and the observed generation times, we generated empirical estimates of the number of secondary infections in the early outbreak stages of the model. We ran 1000 trial simulations from a random single starting infector and quantified i) the mean number of secondary infections from this case, and ii) the time at which each secondary case was infected. We multiplied the rate of infection by a scaling parameter to get a baseline *R*_0_ of 2.8, although we also performed sensitivity analysis (Supplementary Table 1). The mean generation time using this method was 6.3 days (median = 6 days). These basic parameters correspond closely to published estimates^12,30^

In addition to the infection rate from within the network, the infection rate from outside the network is also simulated daily by randomly infecting susceptible individuals with a probability of 0.001 (although we also performed sensitivity analysis of this parameter).

We simulated different contact tracing scenarios using contact information from the network, with the aim of evaluating both app-based and manual contact tracing strategies. Primary and secondary contacts of individuals are identified from the network on the day of the infector’s symptom onset and, as such, contacts of asymptomatic infectors are not traced. Contacts who have already recovered are excluded. Susceptible contacts are traced with a given probability (0.3-0.9 tested - see Supplementary Table 1). We assume that this probability captures a wide range of reasons why contacts might not be traced, and it thus acts as an intuitive simplification.

The isolation and/or quarantine time of each individual is determined based on their infection status, their symptomatic status, whether they have been traced, and the control scenario. We consider four control scenarios: i) no control, where no individuals are isolated or quarantined, ii) case isolation, where individuals isolate upon symptom onset after a delay period, iii) primary contact tracing with quarantine, where individuals isolate upon symptom onset (after a delay) and traced contacts are quarantined upon their infector’s symptom onset (also after a delay), and iv) secondary contact tracing, as scenario iii) but including contacts of contacts. All isolated and quarantined individuals are contained for 14 days.

Finally, we simulated a range of testing efforts for SARS-CoV-2. Each individual is assigned a testing time on isolation or quarantine, with the delay between containment and testing sampled from a Weibull distribution. A cap of the maximum number of daily tests is assigned, and each day up to this number of individuals are randomly selected for testing. Test results are dependent on infection and asymptomatic status, with a false negative rate (i.e. the probability that an infectious case will test negative) 0.1^21,^and a false positive rate (i.e. the probability that susceptible case will test positive) of 0.02^22^ Cases who tested negative were immediately released from isolation/quarantine.

A set of default parameters were chosen to represent a relatively optimistic model of contact tracing, which included a short time delay between symptom onset/tracing and isolation/quarantine (1-2 days), and a high proportion (90%) of contacts traced within this tracked population (default parameters highlighted in bold in Supplementary Table 1). We assumed that the probability of tracing was constant over time, and therefore independent of previous isolation/quarantine events, and that all individuals remained in quarantine for the full 14 days, unless released via testing. We performed sensitivity tests on all relevant parameters (Supplementary Table 1). To examine how infection dynamics were affected by network structure, we ran epidemic simulations on each of the null networks described above. We also ran simulations on networks generated using higher (7m and 16m) distance thresholds for defining a contact. These networks were 20% and 100% more dense, respectively, and therefore provide an estimate of the robustness of our simulations to missing contacts.

We ran each simulation for 70 days, at which point the majority of new infections came from outside the network (see results), with all scenarios replicated 1000 times. With the null networks (above) and physical distancing simulations (below), we ran one replicate simulation on each of 1000 simulated networks. In no simulations were all individuals in the population infected under our default settings. Therefore, for each simulation we report the number of cases per week, and quantify the total number of cases after 70 days as a measure of outbreak severity. To present the level of isolation and quarantine required under different scenarios, we calculate the number of people contained on each day of the outbreak, and average this ove weeks to get weekly changes in the daily rates of isolations and quarantines.

### Physical distancing Simulations

We simulated a population-level physical distancing effort, whereby a given proportion of the ‘weak links’ are removed (edges only observed on a single day; Extended Data Fig. 10A-D). This is akin to a simple situation whereby individuals reduce their non-regular contacts (e.g. to people outside of their household or other frequently visited settings such as workplaces). As further supplementary analysis, we also carried out a more complex physical distancing simulation, whereby the weak links that were removed were randomly reassigned to existing contacts (Extended Data Fig. 10E-G). This represents a scenario where individuals reduce their non-regular contacts but spend more time with regular contacts.

The epidemic model code can be accessed at: https://github.com/biouea/covidhm

## Acknowledgements

This work was instigated through the Royal Society’s Rapid Assistance in Modelling the Pandemic (RAMP) scheme. We thank Michael Pointer for helpful discussions throughout, and Cock van Oosterhout and Julia Gog for comments on the manuscript. We thank all those in Haslemere who took part in the BBC Pandemic study. We thank Hannah Fry and 360 Production, especially Danielle Peck and Cressida Kinnear, for making possible the collection of the dataset that underlies this work, and Andrew Conlan, Maria Tang, and Julia Gog for their contribution to the BBC study. JAF was supported by a research fellowship from Merton College and BBSRC (BB/S009752/1) and acknowledges funding from NERC (NE/S010335/1). PK was in part funded by the Royal Society under award RP\EA\180004 and European Commission: 101003688. AJK was supported by a Sir Henry Dale Fellowship jointly funded by the Wellcome Trust and the Royal Society (grant Number 206250/Z/17/Z).

## Data availability

This study used the raw data previously published in Kissler et al. 2018 (made available with full description here: https://www.biorxiv.org/content/10.1101/479154v1). The data used here are publicly available with the code

## Code availability

The code and data used to produce the simulations is available as an R package at: https://github.com/biouea/covidhm. A shiny app which runs individual outbreak simulations is available at: https://biouea.shinyapps.io/covidhm_shiny/

## Author contributions

J.A.F. A.J.K. and L.G.S. conceived the study; J.A.F. carried out the social network analysis, with input from P.K., S.K., A.J.K and L.G.S; L.G.S. built the epidemic network model with input from J.A.F., J.H., S.K., P.K. and A.J.K; J.A.F. and L.G.S. wrote the first draft of the manuscript; All authors interpreted the results, contributed to writing and approved the final version for submission.

